# Biobehavioral pain profiling of minoritized adults with chronic widespread pain and clinical obesity before and after bariatric surgery: study protocol for a longitudinal, observational cohort study

**DOI:** 10.64898/2026.08.04.26359660

**Authors:** Ericka N Merriwether, Mariam Maqsood, Sally M Vanegas, Steven Em, Natalia Perez, Manish Parikh, Blanca Ruiz-Guerenabarrena, Claudia Humala-Martinez, Briana Lopez, Roger B. Fillingim, Melanie Jay

**Affiliations:** Department of Physical Therapy, NYU Steinhardt School of Education, Culture, and Human Development, New York University, New York, NY, USA; Department of Medicine, NYU Grossman School of Medicine, New York University, New York, NY, USA; New York City Health and Hospitals/Bellevue Hospital Center, New York, NY, USA; Department of Surgery, NYU Grossman School of Medicine, New York University, New York, NY, USA; R&R Surgical Institute, Torrance, CA, USA; Pain Research and Innovation Center of Excellence, University of Florida, Gainesville, FL, USA; College of Dentistry, University of Florida, Gainesville, FL, USA; Department of Population Health, NYU Grossman School of Medicine, New York University, New York, NY, USA; New York Harbor Veterans Affairs, New York, NY, USA

**Author notes:** **Corresponding Author:** Dr. Ericka N Merriwether, PT, DPT, PhD, NYU Steinhardt Department of Physical Therapy, NYU Grossman School of Medicine, 380 Second Avenue, 4^th^ Floor, New York, NY, 10010.

**Keywords:** chronic pain, widespread pain, obesity, weight loss, bariatric surgery, function, movement, pain disparities

## Abstract

Chronic widespread pain (CWP) is highly prevalent among minoritized adults with clinical obesity, and symptom management is challenging. Weight loss via bariatric surgery is often recommended to improve musculoskeletal pain. However, there is significant variation in pain trajectories following bariatric surgery, and the impact of weight loss on movement-evoked pain is largely unknown. The current study aims to systematically characterize and quantify longitudinal changes in pain at rest and movement-evoked pain up to 6 months post-surgery, and to determine whether pain modulatory mechanisms, joint motion, and mechanical loading biosignatures mediate the relationship between weight loss and pain change. This study protocol details the research methodologies and procedures for a prospective observational cohort study of 60 individuals undergoing bariatric surgery for weight loss. Participants will complete questionnaires, anthropometric measurements, clinical and experimental pain testing, functional testing, and a standardized movement testing battery to assess joint motion and mechanical loading using camera-based motion capture before and at 3 and 6 months post-bariatric surgery. Generalized linear mixed models to assess the significance of changes in PAR, MEP, and all patient-reported outcome measures. Reduced models will treat the main effect of time as a fixed factor, and intra-individual repeated measures as random effects. Ethics and dissemination: This study protocol has been registered as an observational study with ClinicalTrials.gov (NCT0675386), version #1, in the United States and has been approved by the NYU Langone Health Institutional Review Board (IRB#: i21-01652) and the New York City Health + Hospitals/Bellevue Research Office (Bellevue Study ID #: STUDY00003739). Study results will be published in peer-reviewed journals and presented at national and international conferences and community events.

## Introduction

Chronic pain and obesity are interrelated, highly prevalent conditions that disproportionately affect minoritized populations. Racial disparities in pain burden and restricted access to effective pain management persist(1). Concomitantly, non-Hispanic Black/African- American (NHB) and Hispanic/Latino/a/X/e adults have obesity rates of 40% or higher in the United States(2). Globally, over 2 billion adults are classified as overweight or having preclinical or clinical obesity (BMI > 30)(3), with an estimated 3.8 billion projected by 2050(4). The global economic burden of musculoskeletal conditions associated with higher BMI amounts to approximately $180.7 billion, affecting both high- and low-income countries, though regional differences exist(5). Furthermore, weight bias and perceived healthcare discrimination contribute to pain mismanagement(6). These issues, along with healthcare provider biases about pain reports(7, 8), frequently worsen pain for people with higher BMI or clinical obesity and lead to costly delays in seeking care(9). Therefore, understanding mechanisms of disparity in chronic musculoskeletal pain among minoritized adults with clinical obesity and higher BMI is crucial to the advancement of pain and obesity research, the promotion of health justice, and the mitigation of the global economic healthcare burden of pain.

Chronic widespread pain (CWP) is a subtype of persistent primary musculoskeletal pain (ICD-11 code)(10) characterized by pain at multiple anatomical sites, involving at least four regions and three quadrants of the body, lasting for at least three months(11, 12). CWP is a core symptom of regional and chronic overlapping pain conditions like knee osteoarthritis, persistent non-specific low back pain, and fibromyalgia(13). There are racial and gender-based disparities in CWP, specifically fibromyalgia syndrome (FMS), that are likely a consequence of underdiagnosis despite meeting ACR 2016 diagnostic criteria and greater symptom severity (14). Managing CWP is notoriously challenging, and recommended management strategies often include a combination of multimodal non-pharmacological interventions (15–17). However, there is a notable underrepresentation of minoritized adults with CWP conditions in clinical trials(18). Movement-evoked pain (MEP), musculoskeletal pain experienced during active and passive movement, is more intense than pain at rest (PAR)(19, 20). Moreover, MEP is a key driver of disability in NHB adults(21). Importantly, body size and clinical obesity influence the chronicity and progression of widespread musculoskeletal pain(22). As a result, weight loss is frequently recommended for CWP.

Weight loss is recommended to manage preclinical and clinical obesity and to improve musculoskeletal pain and overall health. However, achieving and maintaining clinically meaningful weight loss through lifestyle interventions alone can be difficult(23). Recent studies have shown that modest improvements in resting pain intensity or disability can be achieved with behavioral lifestyle weight management interventions for adults with knee or hip osteoarthritis or chronic low back pain, though the results are controversial(24, 25). A total weight loss of at least 20% is required to achieve clinically significant improvements in knee pain and physical function (26, 27). Notably, the role of weight and size in MEP characteristics is largely unknown. Obesity care medications (OCMs), such as glucagon-like peptide 1 (GLP-1) receptor agonists (e.g., semaglutide or tirzepatide – Wegovy, Mounjaro), induce clinically significant weight loss and improvements in musculoskeletal pain(28). However, OCMs are costly and require long-term use to maintain weight. Moreover, minoritized adults are less likely to be prescribed OCMs(29), and long-term adherence and health outcomes are still being clarified(30–32). Bariatric surgery is an effective weight-loss intervention for the remission of obesity and other health conditions(32, 33). Importantly, a greater number of NHB and Hispanic/Latino/a/X/e adults are choosing to undergo bariatric surgery than in previous years for the mitigation of localized musculoskeletal pain and overall health improvement(33). However, limited racial and ethnic diversity in weight loss studies that examine pain significantly hinders the longitudinal monitoring of CWP disparities and pain trajectories. Recent work has shown there are racial weight loss disparities(34, 35) and related differences in pain trajectories after bariatric surgery(22). However, the mechanisms underlying these disparities and the short-term pain responses to weight changes need to be elucidated. Thus, a longitudinal observational study of how biobehavioral CWP mechanisms respond to weight loss will significantly advance our understanding of chronic pain phenotypes and their associated states and traits (36) in the context of clinical obesity.

### Study Objectives

The primary aims of this study are to systematically characterize and quantify longitudinal changes in pain at rest (primary outcome) and MEP (secondary outcome) up to 6 months following bariatric surgery. Additionally, the study aims to quantify longitudinal changes in pain modulation and joint movement biosignatures at 6 months post-bariatric surgery. An exploratory, tertiary aim is to determine whether pain modulatory mechanisms, joint motion, and mechanical loading biosignatures mediate the relationship between weight loss and changes in pain. The working hypotheses for this study are: 1) pain at rest and MEP will demonstrate clinically significant reductions post-bariatric surgery; 2) nociceptive processing and joint biomechanics will improve following surgery; and 3) modifications in nociceptive processing and joint movement biosignatures will mediate the association between weight loss and pain reduction. The following narrative describes the study protocol.

## Materials and methods

All information is presented in accordance with the Standard Protocol Items: Recommendations for Interventional Trials – Outcomes 2025 Checklist(37–39). The study is registered with ClinicalTrials.gov (NCT0675386).

### Patient and public involvement

Patient advocates on the NYU Langone Clinical and Translational Science Institute (CTSI) Patient Advisory Council for Research (PACR) were directly consulted on study design, recruitment strategies, and compensation structure for study participation prior to data collection. Patients who were eligible for bariatric surgery and who also had chronic widespread pain were not directly consulted on study design and research methodologies.

### Study Design

The study will employ a prospective, quasi-experimental observational cohort study design.

### Study Setting

Prospective participants will be recruited from two bariatric surgery and weight management clinical sites in New York City, NY, USA. Patients will be recruited in person and by telephone from New York City Health + Hospitals/Bellevue Hospital Center (NYC H+H/Bellevue) and NYU Langone’s Weight Management Program. Prospective study participants will be recruited during bariatric surgery scheduling. Study recruitment began on August 1, 2023. The projected end of the recruitment period will be March 1, 2028. Recruitment and data collection are ongoing. The NYC H+H/Bellevue Center for Obesity & Weight Management is a multidisciplinary program that offers comprehensive weight management services within a publicly funded academic medical center. Patients receive a suite of medical, surgical, and lifestyle modification weight management options in consultation with primary care physicians, psychologists, nutritionists, and bariatric surgeons, and will continue to receive this standard of care throughout the study. The research study team will include clinician-scientists (2 MDs, 1 physical therapist, 2 PhD research trainees who are physical therapists, 2 research assistants), biostatisticians, and 4 research consultants with expertise in weight management, obesity science, and pain neuroscience. The research assistants will be trained by the physical therapist in all data collection methods, and inter-rater reliability will be determined as part of the training protocol. The research team members who will recruit and obtain informed consent are fluent in conversational English and Spanish.

### Human Research Protections

The study has been approved by the NYU Langone Health Institutional Review Board (IRB # i21-01652) and the New York City Health and Hospitals/Bellevue Hospital Center Office of Research (Bellevue Study ID #: STUDY00003739). All research study personnel have completed human subjects research training modules for NYU Langone Health and NYC H+H/Bellevue Hospital Center before recruitment or data collection activities. Research personnel will report unanticipated problems and adverse events to the bariatric surgeon of record, the NYU Langone IRB, and the NYC H+H/Bellevue Hospital Center Office of Research.

### Participant Eligibility Criteria and Recruitment

Written informed consent will be obtained in English or Spanish at the NYC H+H/Bellevue Bariatric Surgery Clinic (primary site) and NYU Langone Tisch Bariatric Clinic (secondary site) prior to study enrollment.

#### Study Eligibility Criteria

Study participants will be new patients from the NYC H+H/Bellevue or NYU Tisch Weight Management Clinics. We will recruit participants who self-identify as Hispanic/Latino/a/X/e and/or NHB based on the 2020 U.S. Census racial categories(40). Demographic information will be collected using the NIH PhenX Toolkit Common Data Elements(41). Participants can select nationality and cultural affiliation, country of origin, gender identity, sex assigned at birth, sexual orientation, current address, household income, and educational level.

Inclusion criteria for study participation are adults 18 to 75 years of age who have elected to undergo bariatric surgery, have pain of at least 3 out of 10 on the Brief Pain Inventory (BPI) Severity subscale at 3 or more anatomical sites, are literate and numeracy fluent in conversational English or Spanish, and have the cognitive capacity to consent to study participation. The main study exclusion criteria are patients who either have pain at less than 3 anatomical sites or pain intensity less than 3 out of 10 on the BPI upon screening, are undergoing revision of a previous bariatric surgery, have localized, acute pain within 6 weeks of study enrollment, or are unable to walk a minimum of 10 feet (3.05 m) unassisted or using a single point cane.

Study eligibility will be determined at 1 of 3 phases of study recruitment. Participants will be approached by a member of the research study team during their initial visit to both study sites and asked if their preferred conversational language is English or Spanish. Research study personnel will then briefly introduce participants to the study. Participants will be informed that their participation in the study is strictly voluntary and will not affect the standard of care before or after bariatric surgery. They will also be informed that they are not financially responsible for any data-collection costs and will be compensated for their participation. If the patient agrees, a screening questionnaire will be administered to participants in English or Spanish. If the patient is eligible for study participation, a research team member will contact them by telephone to explain the study protocol in greater detail and to schedule the first of three data collection sessions (Visit 1). Research personnel will invite participants to meet with the principal investigator to address additional questions or concerns in English or Spanish.

Participants will be enrolled in the study after obtaining written informed consent in English or Spanish. Consent will be obtained in person or remotely using a secure electronic platform. After obtaining written informed consent, study participants will be further screened in person at Visit 1 for neurological, sensory, or motor impairments not assessed during the first 2 phases. If participants demonstrate neurological, sensory, or motor impairments during this examination, they will be deemed ineligible to continue in the study.

### Participant Protections

This is a prospective, quasi-experimental observational study, and all participants will receive standard-of-care bariatric surgery services throughout the study period (42). Study participants will be contacted by telephone, text, and email within 48 hours of data collection to monitor for adverse events associated with pain and movement tests that may elicit discomfort. Clinical pain will be assessed 3 months after bariatric surgery via surveys and telephone contact with participants to gather information on surgery type, current medications, and other relevant health-related issues. An independent study advisory board that is independent from the funder or sponsor will review data collection procedures and evaluate reports of study-related adverse events, such as a worsening of clinical pain or physical function, provided by study personnel. A data monitoring committee appointed by the funder or sponsor is not required because the current study is not a randomized clinical trial. Patient confidentiality will be ensured through multiple methods, including, but not limited to, study ID numbers on forms containing protected health information (PHI) or protected identifying information (PII), and data storage on encrypted servers.

### Sample Size Determination

The sample size needed to achieve the study’s primary aim was determined using extant clinical data from a longitudinal observational cohort study of surgical weight loss(43) (NYU IRB# 16-01995). The primary objective of the study is to characterize and quantify changes in pain at rest (PAR, primary outcome) and MEP (secondary outcome). Power analyses were conducted based on the anticipated within-person mean change in pain at rest from baseline to 6 months post-surgery (primary outcome). Additionally, the clinical assumptions supporting our sample size determinations were the minimum clinically important difference (MCID) for the Brief Pain Inventory (BPI) average pain (2.1 points) and BPI Severity (2.2 points) scores in adults with fibromyalgia (44). The sample size was calculated to detect a Cohen’s d of 0.50 for BPI Severity scores at 3 months, based on a secondary analysis of the parent study cohort, which revealed a sharp reduction in pain intensity ratings in a stratified cohort of NHB and/or Hispanic/Latino/a/X/e adults(22). We considered a 1-point reduction in pain interference at 6 months to be conservative, as proposed in the Initiative on Methods, Measurement, and Pain Assessment in Clinical Trials (45). The sample size calculation assumed a Type I error rate of 2.5% to account for multiple comparisons and 90% power. We estimated a sample size of 50 using a two-sided paired t-test. Thus, we will recruit at least 60 participants to account for a 20% attrition rate, although attrition in the target study population may be higher(46).

### Data Collection

#### Overview (Figure 1)

We will assess specific core and recommended clinical outcomes for weight management interventions(47). There will be two in-person study visits and one remote study visit. Participants will be remunerated $240 for their time, with payments disbursed in two installments. The first payment will be disbursed immediately after completion of Visit 1 and the second after Visit 3. After obtaining written consent during the first visit, data collection will begin with measurements of blood pressure, skin temperature, and oxygen saturation (pulse oximetry). Pain and fatigue at rest will be evaluated, followed by anthropometric measurements and a brief screening for large-fiber peripheral neuropathy. Participants will then undergo clinical assessments of bilateral knee and lumbar spine range of motion using goniometry and gravity-dependent inclinometry, and quadriceps strength will be measured with hand-held dynamometry. Next, a standardized quantitative sensory testing battery will be administered to evaluate pain sensitivity using pressure pain threshold (PPT), mechanical temporal summation (TS), cold tolerance via the cold pressor test, and descending pain inhibition through conditioned pain modulation (CPM). Following this, participants will complete a series of questionnaires in either English or Spanish prior to the administration of performance-based assessments of physical function. Finally, participants will perform a standardized series of evocative movement tasks, which will be recorded using camera-based motion capture (stereophotogrammetry). At the first follow-up visit (3 months), participants will complete pain questionnaires and health surveys either via the REDCap secure online platform or by telephone with a research staff member. During the second follow-up visit at 6 months (Visit 3), the tests and measures completed during the first study visit will be repeated. The anticipated time to complete Visits 1 and 3 will be 2.5 – 3.0 hours, and 0.5 – 1.0 hours for Visit 2. Table 2 provides a detailed summary of the data collection protocol and study timeline.

**Figure 1.**
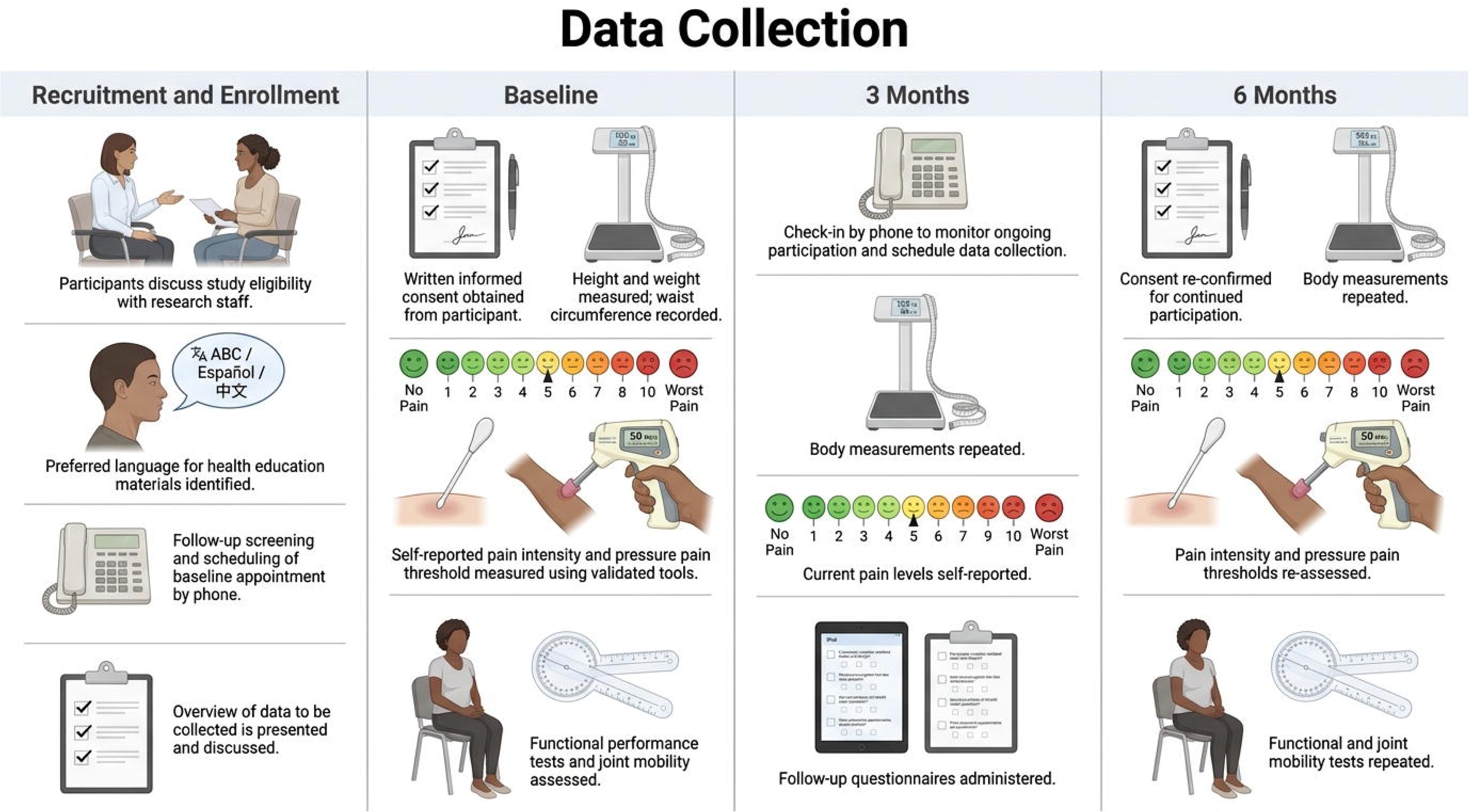
SPIRIT 2025 Checklist.

**Figure 2.**
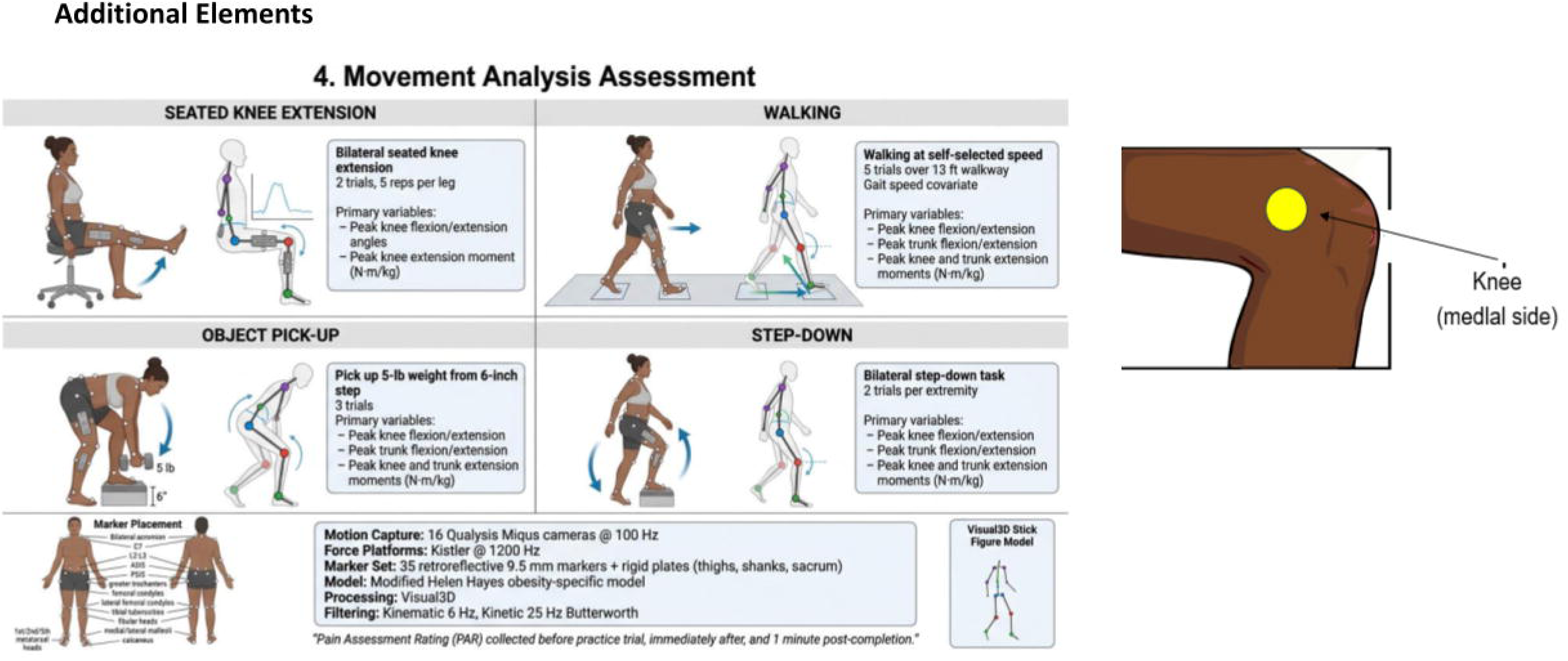
Summary of Data Collection Tests & Measures.

**Table 2. Data collection timeline.**

Baseline (B): prior to bariatric surgery; 3M: 3 months after bariatric surgery; 6M: 6 months after bariatric surgery. PROMIS: Patient-Reported Outcomes Measurement Information System

### Data Management and Security

All demographic, vital signs, anthropometric, pain, physical function, and health-related data will be entered into the Research Electronic Data Capture tool (REDCap). The Research Electronic Data Capture (REDCap) tool, hosted by NYU CTSI, will be used for study data collection and management(48, 49). REDCap is a validated web-based software platform that supports data entry, auditing, and tools for data export and integration. Data will also be manually entered on hard copies of data collection documents to ensure data fidelity. Data standards will be employed when using the NIH Common Data Elements Repository (CDE), the Phenotypes and Exposures (PhenX) Toolkit (https://www.phenxtoolkit.org/), and the Patient-Reported Outcomes Measurement Information System (PROMIS)(50). A bilingual study team member fluent in English and Spanish will administer all surveys and questionnaires. Data will be collected at baseline (Visit 1), 3 months after bariatric surgery (Visit 2), and 6 months after bariatric surgery (Visit 3), the primary study endpoint. Upon request, participants can complete hard copies of surveys or complete them in person at Visit 1 and 6 months post-surgery (Visit 3). For Visit 2, participants can complete digital surveys using the secure REDCap tool or review surveys with bilingual research personnel via telephone or a secure HIPAA-compliant web conferencing platform. All study data will be managed and shared in accordance with the principles outlined in the HHS regulatory guidelines for the protection of human subjects (45 CFR 46). Data will be de-identified using the participants’ assigned study identification number. Study data will be stored for 3 years after the last participant completes the last study visit or for 5 years after the final manuscript is accepted for publication. Study monitoring will occur in 6- month intervals to assess recruitment progress and for interim analyses, primarily descriptive statistics and data quality assessment.

### Anthropometric Measurement and Health Status

Anthropometric and health conditions will be collected at all time points, guided by the bariatric surgery Core Registry Set (CRS)(51) and the Standardised Reporting of Lifestyle Weight Management Interventions to Aid Evaluation (STAR-LITE)(47).

#### Anthropometric measurement

Standardized BMI and waist circumference measurements will be collected using the PhenX BMI Toolkit (https://www.phenxtoolkit.org/ PhenX Toolkit. (2025 September 06). Waist Circumference - Waist Circumference NCFS. https://www.phenxtoolkit.org/protocols/view/21602). Body weight (kg) will be measured in light clothing, without shoes, using a Stow-A-Weigh digital bariatric hospital scale (Scale-Tronix, Welch Allyn, Skaneateles, NY, USA). Height (cm) will be measured using a stadiometer (SECA 213, Seca GmBH & Co. KG, Hamburg, Germany). BMI will be calculated using anthropometric measurements (kg/m^2^) and categorized according to the World Health Organization: Class I=30- 34.99 kg/m^2^, Class II=35-39.99 kg/m^2^, and Class III=40 kg/m^2^ or higher. Waist circumference measurement is a proxy for central adiposity and is associated with an elevated risk of Type 1 (52) and Type 2 diabetes, dyslipidemia, and cardiovascular disease (53). It will be measured at the midpoint between the iliac crest and the lowest rib (between the 10th and 12^th^ ribs)(54). The umbilicus will be used as an additional anatomical reference point to improve measurement accuracy. The horizontal distance between the left and right anterior superior iliac spines (inter-ASIS distance) will be measured using a tape measure(55). The inter-ASIS distance will be used to assist in biomechanical modeling of the pelvis and lower-extremity segments’ position and pose during motion analysis (56). The percentage of total weight loss (%TWL) will be the weight loss outcome variable. The %TWL is the ratio of weight loss after surgery to presurgical weight, calculated as (baseline weight − current weight) / (baseline weight) × 100 (57). The percentage of TWL will be calculated at 6 months after bariatric surgery.

#### Sensory Testing

Participants will undergo sensation testing at select sites on the bilateral lower extremities while seated in a chair at standard height with arm support. Light-touch sensation and allodynia will be assessed by applying a cotton ball to the L4-S1 dermatomal distribution over the lower leg, ankle, and foot. Mechanical cutaneous sensation will be assessed using a 5.07 g Semmes-Weinstein monofilament on the foot at the following sites: dorsal surface, between the 2^nd^ and 3^rd^ metatarsals, and the plantar surface of the 1^st^, 3^rd^, and 5^th^ metatarsal heads. The order of application of the cotton ball and monofilament across anatomical sites will be randomized.

#### Health status

Surgical intervention type (Roux–en–Y, sleeve gastrectomy, or Lap-Band), medical comorbidities (e.g., diabetes, cardiovascular health), and current medications for pain and other health conditions will be obtained from participants’ electronic medical records.

### Pain Measurement and Assessment

#### Pain Intensity at Rest (PAR)

Pain intensity at rest (PAR) will be evaluated using the Numeric Pain Rating Scale (NPRS) and the Brief Pain Inventory – Short Form (BPI-SF).

#### Numeric Pain Rating Scale (NPRS)

The NPRS is a single-item pain intensity scale ranging from 0 (no pain) to 10 (worst pain imaginable). The NPRS has been validated in Spanish-speaking populations(58).

#### Brief Pain Inventory – Short Form (BPI – SF)

The BPI-SF is a 9-item self-assessment that measures pain intensity, interference with physical functioning, and pain location(59). The BPI-SF Pain Severity and Pain Interference subscales feature anchors from 0 (“no pain”) to 10 (“Pain As Bad As You Can Imagine”), and the subscale score is the mean of the four items. The BPI-SF Pain Interference subscale is the mean of 7 items. The BPI-SF has been validated in Spanish (60) and in adults undergoing bariatric surgery (61, 62). The BPI-SF assesses pain domains recommended by the Initiative on Methods, Measurement, and Pain Assessment in Clinical Trials (IMMPACT)(45, 63). Pain at rest will be measured before vital signs and anthropometric measurements are taken.

#### Movement-Evoked Pain (MEP)

MEP intensity will be a secondary outcome of this study and will be calculated as a composite score of pain intensity ratings across a battery of functional movement tasks (64). MEP will be measured during objective measures of physical function (5TSTS, 6MWT) and during a standardized battery of 4 evocative movement tasks: seated knee extensions, walking, picking up a 5 lb. (2.27 kg) weight, and ascending and descending a 6-inch step. More detailed descriptions of each movement task are provided below. During the 6MWT, participants will rate their pain intensity using the NPRS (0 = no pain, 10 = worst pain imaginable) before the test.

Participants will rate their pain at the 5-minute mark of the test and again immediately after the 6MWT. During the 5TSTS, participants will rate their pain immediately and 60 seconds after the last repetition.

Pain intensity ratings and location will be obtained immediately following and 30 seconds after evocative pain tasks using the NPRS for the standardized movement testing battery. The MEP composite score will be calculated as the average pain intensity rating across all movement tasks, divided by the number of movement tasks. MEP aggregate scores will be calculated separately for objective functional measurements and for the standardized evocative task movement testing battery. Higher scores indicate greater MEP.

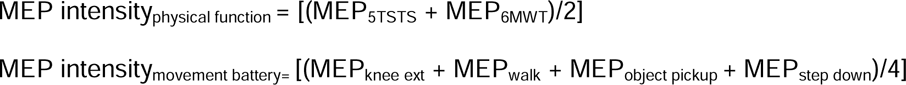

#### Pain Location and Chronicity

The number and location of painful anatomical sites will be assessed using the BPI-SF. Participants will complete Question 2 of the BPI-SF, which features a numerically coded body diagram showing 45 painful sites in the coronal (mediolateral) plane. Participants will self-report the duration of their chronic pain in months or years.

#### Pain Interference

In addition to the BPI, pain interference will be assessed using the Patient-Reported Measurement Information System (PROMIS) Pain Interference Short-Form 8a. The PROMIS Pain Interference Short-Form 8a assesses the impact of pain on social and recreational activities (65). Items are rated using a Likert scale and summed to compute a total raw score. Raw scores are then converted to a standardized T-score, with a mean of 50 for the general population. Higher T-scores (>50) indicate greater pain interference. The PROMIS Pain Interference Short–Form 8a has been validated in chronic pain populations (66, 67) and is available in Spanish (68).

#### Pain Behavior

Behaviors indicating that a person has experienced pain in the last 7 days will be assessed using the PROMIS SF v1.0 - Pain Behavior Short-Form 7a (69). Items are rated on a 6-point Likert scale, with responses ranging from “Had no pain” to “Always”. Items are summed to yield a total raw score, which is then converted to a standardized T-score. Higher T-scores (>50) indicate more pain behaviors. The PROMIS Pain Behavior Short-Form 7a has been validated in women with fibromyalgia(70).

#### Central Sensitization

The Central Sensitization Inventory (CSI) is a 25-item survey that queries symptoms indicative of central sensitization (71). The items ask how frequently respondents experience symptoms on a Likert scale with anchors from 0 (never) to 4 (always). The CSI total score ranges from 0 to 100, with higher scores indicating greater central sensitization. A score of 40 or higher yielded good sensitivity and specificity for detecting central sensitization(72). The CSI has been validated in multilingual populations with chronic pain(73).

### Biobehavioral Experimental Pain Assays

#### Pressure Pain Threshold

Mechanical pressure pain threshold (PPT) will be measured using a manual digital pressure algometer (Somedic AB, Farsta, Sweden) to assess both localized and widespread static pain sensitivity by activating Group III and Group IV muscle nociceptors (deep-tissue muscle afferents) (74, 75). Mechanical pressure will be applied using a 1cm^2^ algometer probe at a ramp rate of 50 kPa/sec. To ensure safety, the applied pressure will not exceed 700 kPa/sec. The three anatomical PPT testing sites are the muscle belly of the upper trapezius at the midpoint between the C7 vertebral spinous process and the acromion, the paraspinal muscles 1-2 cm lateral to the intervertebral space between the L2/L3 spinous processes (lower back)(76), and the medial knee joint line, the intercondylar space between the medial femoral epicondyle and the medial tibial condyle (21). These regions have previously been validated in adults with widespread pain (75). Pressure measurements will stop and be recorded once the participants press and release an external button connected to the device. Scripted participant instructions will be: “You will feel pressure at first. When you start to feel pain around 1 out of ten on the pain scale, press and release the red button.” These scripted instructions will also be given in conversational Spanish. There will be three repetitions at each anatomical testing site, and the mean PPT value in kilopascals (kPa) for each site will be used for all analyses. Lower PPT values indicate greater static pain sensitivity in deep-tissue muscle afferents. Details are outlined in **Supplementary Figure 1**.

#### Cold Pressor Test (CPT)

We will also measure responses to noxious cold stimuli using a modified cold-pressor test (CPT)(77). Participants will be asked to submerge their non-dominant hand up to the wrist crease in a mixture of ice and water in a 10-liter (2.5-gallon) plastic pail. The water temperature will range from 8 °C to 12 °C, with a test duration of 2 minutes or until the participant reaches maximum tolerable pain to ensure participant safety. Participants will rate their pain intensity on a scale from 0 to 10 during and 30 seconds after hand immersion. During the CPT, we will also measure conditioned pain modulation (CPM), as outlined below. Cold pain intensity ratings and immersion time will be recorded and used in the analyses.

#### Mechanical Temporal Summation (TS)

Mechanical temporal summation (TS) will be conducted with a predetermined stimulus intensity of 256 mN using a weighted pinprick stimulator (MRC Systems GmbH, Germany) to assess dynamic pain sensitivity. TS measurements will be taken at the intervertebral space between the L2/L3 spinous processes (lower back), which is the primary anatomical site of pain, as our previous work showed a higher prevalence of low back pain among racialized patients undergoing bariatric surgery (22). TS will also be measured at two remote anatomical sites: at the thenar eminence and the middle phalanx on the dorsum of the non-dominant hand (78). Pilot testing showed that 256 mN was best tolerated and elicited a summation response (data not shown). The anatomical location of the TS measurement was determined from prior literature on racialized Hispanic/Latino/a/X/e participants (79). Participants will be familiarized with the test using a single-probe application to the dominant forearm. The test will begin with a single pinprick stimulus, during which participants will verbally rate their pain intensity. There will be a single 10-train stimulus application, with the probe applied perpendicularly to the anatomical site at 1 Hz (1 cycle per second). Participants will be asked to verbally rate their pain immediately and 30 seconds after the 10^th^ stimulus. The absolute effect of TS will be calculated as the difference in pain ratings between the 1^st^ stimulus and the 10^th^ application at each anatomical site. A larger absolute effect of TS indicates greater dynamic pain sensitivity. Details are outlined in **Supplementary Figure 2**.

#### Conditioned Pain Modulation (CPM)

We will perform conditioned pain modulation (CPM) to assess the function of descending inhibitory pathways responsible for endogenous analgesia (80). The total duration of CPM testing will be 2 minutes. We will use the parallel CPM paradigm, which has been shown to elicit a more robust inhibitory response in women with fibromyalgia (81). The test stimulus will be mechanical pressure applied at the upper trapezius using a pressure algometer contralateral to the hand submerged in cold water. The conditioning stimulus will be cold-water immersion of the non-dominant hand up to the wrist in an 8 °- 12 °C water bath. PPT will be measured immediately before (PPT1) and 60 seconds after (PPT2) cold water immersion. Pain intensity will be assessed at 20, 60, and 120 seconds after cold water immersion. The average of 3 PPT measurements at the upper trapezius will be included in the analyses. The percent change in PPT2-to-PPT1 values (PPT2/PPT1) x 100 and the absolute effect (PPT2 – PPT1) will be calculated. Lower values indicate impaired function of descending inhibitory pathways. This protocol has been used in women with fibromyalgia and higher BMI (82, 83). Details are outlined in **Supplementary Figure 3**.

### Physical Function

Physical function and performance will be objectively measured at baseline (Visit 1) and at 6 months after bariatric surgery (Visit 3) using a modified Five-Time Sit-to-Stand Test (FTSTS) and the Six-Minute Walk Test (6MWT). Blood pressure, oxygen saturation, heart rate (beats per minute), and ratings of pain and fatigue intensity will be measured before and immediately after test completion. Participants will be asked to rate their MEP intensity as previously described and their level of perceived exertion at the 5-minute mark of the test. Perceived exertion will be assessed using the Borg Rate of Perceived Exertion (RPE)(84). During the FTSTS, participants will begin the test by sitting with their arms folded in their laps and their backs against a chair (45.7 cm (18 in) height x 48.3 cm (19 in) depth x 48.3 cm (19 in) width). Research personnel will instruct the participant to stand with the knees fully extended, then sit in a chair for 5 repetitions (85). The test will begin with the 1^st^ repetition and end after the 5^th^. The time required to complete the test, in seconds, will be included in all data analyses. The 5TSTS has been used in adults with fibromyalgia (15) and people who have had bariatric surgery (86). The 6MWT will be performed in accordance with a modified standardized protocol (87) to ensure participant safety and facilitate MEP assessment. Participants will complete the 6MWT in a corridor along a pre-measured 50-foot (15.2 m) walkway. All participants will receive standardized instructions in either English or Spanish before the test. Walking distance will be recorded in meters and included in all data analyses.

Participants will complete self-reported assessments of physical function. The PROMIS Short-Form v2.0 Physical Function 8b is a self-assessment of respondents’ abilities to perform a variety of physical activities using a 5-point Likert scale where responses range from “Without any difficulty” to “Unable to do”(88). Items are summed to yield a total raw score, which is then converted to a standardized T-score. Lower T-scores (<50) indicate worse physical function. The PROMIS Pain Interference Short-Form has been validated in adults with fibromyalgia(70, 89) and is available in Spanish.

The Lower Extremity Functional Scale (LEFS) assesses difficulty performing functional activities involving the lower extremities. Twenty items are rated on a 5-point Likert scale with anchors from 0 (“Extreme Difficulty/Unable to perform activity”) to 4 (“no difficulty”). The summative score for all items is the total score, ranging from 0 to 80. Lower scores indicate poorer lower-extremity function. Construct validity was moderate to good (r = 0.64 to 0.80) for the English version of the LEFS when compared with SF-36 physical function and physical component summary scores, and test–retest reliability (R = 0.86) and internal consistency (Cronbach alpha = 0.96) were excellent for the English version(90). The Spanish-translated version of the LEFS had high test-retest reliability (ICC=0.998, 95% CI: 0.996-0.999), high internal consistency (Cronbach’s α = 0.989), and moderate to high concurrent validity with the SF-36 subscales (r=0.504–0.903)(91).

### Fatigue

Fatigue intensity will be assessed at the beginning of data collection (fatigue at rest) and before and after objective measures of physical function (movement-evoked fatigue). Fatigue intensity will be assessed using an 11-point rating scale ranging from 0 (no fatigue) to 10 (very fatigued). The PROMIS Short Form v1.0 – Fatigue 13a (FACIT-Fatigue) is a self-reported assessment of the frequency, timing, and severity of fatigue as well as the impact of fatigue on physical, cognitive, and social activities over the past 7 days (88). This survey is derived from the original Functional Assessment of Chronic Illness Therapy (FACIT), which was developed to assess health-related quality of life across health conditions (92). Items are rated on a 5-point Likert scale, with responses ranging from “Not at all” to “Very much”. Items are summed to yield a total raw score, which is then converted to a standardized T-score. Higher T-scores (<50) indicate greater fatigue. These measures have been validated in populations with localized and widespread pain (15, 89), but a minimal clinically important change has not yet been established.

### Psychological, Social, and Emotional Functioning

#### Pain Catastrophizing

Pain catastrophizing is a psychological phenomenon characterized by a tendency to amplify the perceived threat of a painful event, experience feelings of helplessness in the midst of pain, and ruminate about the pain before, during, or after a pain episode(93). The Pain Catastrophizing Scale (PCS) is a 13-item self-report instrument that evaluates levels of pain catastrophizing. Items on the PCS assess the extent to which participants experience magnification, rumination, or feelings of helplessness when in pain. Scores range from 0 to 52, with scores above 30 indicating clinically significant degrees of pain catastrophizing. The PCS is valid and reliable across multiple pain populations(94), and has been used in studies of patients who have undergone bariatric surgery(95), as well as in Spanish-speaking patients with fibromyalgia (Cronbach’s alpha = 0.79, ICC = 0.84(96)).

#### Perceived Stress

The Perceived Stress Scale-10 (PSS-10) assesses the level to which different situations are perceived as stressful over the past 30 days(97). Items query how frequently respondents endorse situations as stressful events on a 5-point Likert scale with anchors from 0 (never) to 4 (very often), with summative scores ranging from 0 to 40. Higher scores indicate greater levels of perceived stress. The PSS-10 demonstrates moderate internal consistency (Cronbach’s alpha > 0.70) across English and Spanish translations(97, 98).

#### Depression

Self-reported emotional distress as a result of a depressed mood will be assessed using the PROMIS Short Form v1.0 - Depression 8a questionnaire(99). Items are rated on a 5-point Likert scale, with responses ranging from “Never” to “Always”. Items are summed to yield a total raw score, which is then converted to a standardized T-score. Higher T-scores (<50) indicate higher levels of depression. The PROMIS Short Form v1.0 - Depression 8a questionnaire has been validated in patients who have undergone bariatric surgery (Cronbach’s alpha = 0.94; inter-item correlation (r = 0.69-0.88); item-scale correlations (r = 0.77-0.81))(100) and in racially and ethnically diverse adults(101).

#### Emotional Support

We will assess perceived levels of emotional support (e.g., being cared for, having a person to confide in) with the PROMIS SF v2.0 - Emotional Support 6a questionnaire(88). Items are rated on a 5-point Likert scale, with responses ranging from “Never” to “Always”. Items are summed to yield a total raw score, which is then converted to a standardized T-score. Lower T-scores (<50) indicate lower levels of emotional support. The criterion and construct validity of the PROMIS SF v2.0 - Emotional Support 6a questionnaire for English and Spanish-speaking populations is good(102), although the psychometric properties of the instrument for patients undergoing bariatric surgery have not yet been established.

#### Sleep Quality

We will assess sleep patterns and quality using the Pittsburgh Sleep Quality Index (PSQI). The PSQI survey is a self-reported assessment of sleep quality, latency, duration, disturbances, sleep efficiency, sleep medication use, and daytime functioning over the last month(103). The PSQI has 19 fixed-choice and open-ended items with 7 components. Component scores are derived from ratings on a 4-point Likert scale with anchors from 0 (no difficulty) to 3 (severe difficulty). The summative score for all components is the total score, ranging from 0 to 21. Higher scores greater than 5 indicate poor sleep quality. The PSQI has been validated in patients with fibromyalgia(104), in Spanish-translated versions(105), and for patients who have undergone bariatric surgery(106).

### Joint Motion and Muscle Strength

#### Range of Motion (ROM)

Bilateral active knee and thoracolumbar spinal ROM in the sagittal plane will be measured using a universal long-arm goniometer and gravity-dependent bubble inclinometry. Active knee flexion and extension will be measured in the supine position, with the hip at 0° of extension, 0° of abduction/adduction, and 0° of internal/external rotation using a Baseline® transparent plastic goniometer (Fabrication Enterprises, White Plains, NY, USA). The axis of the universal goniometer will be placed at the lateral femoral epicondyle. The stationary arm will be aligned with the greater trochanter along the proximal femur, and the moving arm will be aligned with the lateral malleolus along the distal tibia. Active flexion and extension of the lumbar spine will be measured using a Baseline® bubble inclinometer (Fabrication Enterprises, White Plains, NY, USA) according to modified guidelines by Waddell et al.(107) The anatomical reference points for placing a bubble inclinometer to measure cervical flexion and extension are the C7 spinous process and the intervertebral space between the L2/L3 spinous processes. Participants’ starting position will be with their arms at their sides and their feet approximately shoulder-width apart(108). Participants will be instructed to bend forward to touch their toes or lean back as far as possible to assess active end-range thoracolumbar flexion and extension.

#### Motion Analysis

Marker-based motion capture will be used to quantify kinematic (joint and segmental motion) and kinetic (mechanical loading) during a standardized battery of evocative movement tasks. We will use marker-based motion capture (stereophotogrammetry) in an instrumented gait analysis laboratory to measure the three-dimensional positions and orientations of the trunk, pelvis, thigh, shank (leg), and foot segments(109). We will use a modified Helen Hayes “obesity-specific” biomechanical model of the trunk, pelvis, and thigh segments(110). The marker set will include 35 spherical 9.5mm retroreflective markers applied to the following anatomical sites: right and left acromion, C7 vertebral spine, L2-L3 intervertebral space, bilateral anterior superior iliac spine, bilateral posterior superior iliac spine, bilateral greater trochanters, bilateral femoral condyles, bilateral lateral femoral condyles, bilateral tibial tuberosities, bilateral fibular heads, bilateral medial malleoli, bilateral malleoli, bilateral first metatarsal heads, bilateral second metatarsal heads, bilateral fifth metatarsal heads, and bilateral calcaneus. Rigid plates consisting of 3 markers will be taped to the thighs, shanks (lower legs), and the sacrum. Kinematic data will be collected at 100 Hz using 16 Qualysis infrared, video recording cameras (Miqus, Qualysis North America, Inc., Buffalo Grove, IL, USA). Kinetic data will be collected using Kistler force platforms (Kistler Instrumente GmbH, St. Joseph, MI, USA) embedded in a walkway at a sampling frequency of 1200 Hz. Three-dimensional coordinates of the retroreflective markers will be pre-processed in Qualysis Tracking Manager^TM^ (Qualysis North America, Inc., Buffalo Grove, IL, USA). Kinematic models will be developed in Visual 3D^TM^ (C- Motion, Germantown, MD, USA, v2024.08.3). Kinematic data will be filtered using a fourth-order, low-pass Butterworth filter with a cutoff of 6 Hz. Kinetic data will be filtered using a fourth-order low-pass Butterworth filter with a cutoff frequency of 25 Hz.

The movement testing battery will include bilateral seated knee extension, overground walking, picking up a weight from a step (object pick-up task), and a bilateral step-down task (**Figure 3**). Participants will be given scripted instructions in either English or Spanish on how to perform each movement task and will complete one practice trial before recording. Participants will perform knee flexion and extension while sitting on a stool for the seated knee extension task. The stool will be adjusted to the participants’ height to maintain 90 degrees at knee flexion with the thighs parallel to the floor. Two trials will consist of 5 repetitions per leg, and the average of each trial will be used in all statistical analyses. For the walking task, participants will perform 5 trials over a 13 ft (396.2 cm) walkway at their self-selected gait speed. Gait speed will be included as a covariate in statistical analyses because it is associated with weight or BMI(111). The average of 3 trials will be used for statistical analyses. For the object pick-up task, participants will pick up a 5-lb (2.27 kg) weight from a 6-inch (15.2 cm) step and return it to its original position. There will be 3 trials of this task, and the average will be included in statistical analyses. For the step-down task, participants will be asked to ascend and descend a 6-inch step with the same lower extremity. There will be 2 trials for each lower extremity, and the average across trials will be included in the statistical analyses. The primary kinematic variables to be analyzed are peak knee flexion and extension, and peak trunk flexion and extension, across all movement tasks. The primary kinetic variables to be analyzed are peak knee and trunk extension moments (N · m/kg). We will include additional kinematic and kinetic variables in our exploratory analyses. Participants will rate their PAR before the practice trial. Upon completion of the last trial of the movement task, participants will be asked to rate their pain immediately and 1 minute after task completion.

**Figure 3.** Motion analysis testing battery. **A.** *Seated knee extension.* Participants will perform 2 trials of 5 repetitions of seated bilateral knee extensions. **B.** *Walking.* Participants will perform 5 trials walking over an instrumented walkway. **C.** *Object Pick-Up*. Participants will perform 3 trials of picking up a 5 lb. (2.27 kg) weight from a step (15.2 cm) on an embedded force plate. **D.** *Step-Down.* Participants will perform 2 trials per lower extremity of ascending and descending one step (15.2 cm). **E.** Marker placement and biomechanical modeling parameters.

#### Muscle Strength

Bilateral quadriceps strength will be measured using handheld dynamometry (HOGGANS Scientific, LLC, Salt Lake City, UT, USA). The test position will be with participants seated, knees at 90 degrees of flexion. The dynamometer will be placed approximately 1-3 cm proximal to the medial and lateral malleoli, and the tester will administer the “break test,” in which a perpendicular force is applied at the end of the knee extension movement(112). Participants will be instructed to extend their knee and maintain that position against resistance for a 5-second hold(113). There will be two trials per leg, and the average across trials will be used for analysis.

#### Harms and Adverse Events

Research personnel directly involved in data collection will be responsible for managing and reporting study harms and adverse events. Serious adverse events are not anticipated, and the study underwent an Expedited Review as it met the criteria for minimal risk. Unanticipated harms and adverse events will be queried and non-systematically documented electronically or by telephone within 72 hours after data collection and reported by authorized research personnel. Classification and grading of the severity of harms and adverse events will be documented by the research personnel directly involved in data collection, with additional review and final classification by the Principal Investigator. All unanticipated harms and adverse events that occur during data collection will be documented in REDCap at the time of occurrence and reported to the IRB within 7 business days. Continued study participation after an unanticipated harm or adverse event will be determined by institutional guidance, clinical decision-making, and participants’ decisions to continue or withdraw from the study.

### Planned Statistical Analyses

Participant demographics, anthropometric data, clinical pain and movement data, and patient-reported outcomes will be reported for the entire study cohort at baseline and at 6 months post-surgery. We will use frequency analyses to determine the point prevalence of widespread pain (i.e., relative frequency) within the cohort at all time points. The proportion of participants who meet the CWP inclusion criteria at baseline will be calculated. We will use log transformations or arc-sin-square-root transformations, as appropriate, for proportion data. We will also evaluate whether changes in PAR are increasing, decreasing, or stable between baseline and 6 months post-surgery using trend analyses on record-level data. We will also conduct an initial repeated analysis of covariance (ANCOVA).

#### Primary and Secondary Aims

The study’s primary aim is to systematically characterize and quantify changes in PAR 6 months post-surgery. The secondary aim is to quantify changes in MEP at the same study endpoint. NPRS pain ratings will be averaged, and change scores (PAR _baseline_ – PAR _6M_) will be calculated at baseline and 6M post-surgery (primary study endpoint). Aggregate MEP scores will be analyzed at baseline and at 6 months post-surgery. Since data will be correlated due to repeated measures of pain-related outcomes within the same participant, we will apply mixed-effects models at the record level to evaluate longitudinal changes in PAR after bariatric surgery(114). This model accounts for differences in PAR slope before and after a specified time event. Previous studies from our group indicate differences in PAR slope between baseline and 3 months post-bariatric surgery, and between 3M and 24 months post-surgery(115).

### Exploratory Aim

To determine whether the percentage of total weight loss (%TWL) is associated with changes in pain, we will initially use bivariate correlations. We will then employ generalized linear mixed models to assess the significance of changes in PAR, MEP, and all patient-reported outcome measures. We will examine the differences in the primary, secondary, and exploratory outcomes between NHB and Hispanic/Latino/a/X/e participants using piecewise linear mixed-effects models. Reduced models will treat the main effect of time as a fixed factor, and intra-individual repeated measures as random effects. In the full models, adjustments will be made for Time, Age, Race, and Gender. We will conduct pairwise comparisons to determine differences in estimates of pain-related outcomes between the two racial groups at each time point, using the Benjamini-Hochberg procedure for multiple comparisons (116). Statistical significance will be set at p > 0.005 for a two-tailed test, accounting for multiple comparisons.

### Sensitivity Analyses and Missing Data

To evaluate how biases and assumptions might influence the study results, we will perform pre-planned sensitivity analyses. Data will be examined visually and statistically for normality using Q-Q plots and Shapiro-Wilk tests. Data quality will be monitored throughout the study to detect patterns of missingness among primary, secondary, and exploratory variables. Missing data patterns will be analyzed using the appropriate statistical tests based on the variance between and within imputations relative to the total variance. If data are found to be completely missing at random, we will use multiple imputation methods to assess the impact of missing data on our analyses. We will report the number and percentage of missing values for all variable types. Attrition bias will be evaluated by comparing participants with and without baseline missing data to identify patterns in demographic and other variables. Additionally, we will ask participants for their reasons for withdrawal, if possible. To analyze subgroup differences in primary, secondary, and exploratory variables across multiple time points, we will use the False Discovery Rate (FDR) correction and the Benjamini-Hochberg procedure to control for multiple comparisons(116).

#### Ethics and Dissemination

The study was approved by the NYU Langone Health Institutional Review Board (Study #: i21-01652) and by NYC Health + Hospitals/Bellevue Office of Research (Bellevue Study ID #: STUDY00003739). All study procedures will be conducted according to principles outlined in the Declaration of Helsinki and in the International Council for Harmonisation (ICH) Integrated Addendum to ICH (R3) Guideline for Good Clinical Practice(117), as well as other institutional and municipal regulations. The study will undergo annual reviews and auditing upon request to ensure safety and compliance with federal regulations. All protocol amendments will be subject to approval by the NYU Langone Health Institutional Review Board and NYC Health + Hospitals/Bellevue Office of Research. Substantive study modifications will be communicated to currently enrolled participants in English or Spanish, and updates to trial registries will be made within 30 days of institutional approval.

Findings will be reported in accordance with the Strengthening the Reporting of Observational Studies in Epidemiology (STROBE) Guidelines for observational studies and the principles outlined in the Guiding Principles for Equity in Reporting (118, 119). Authorship eligibility will be determined in accordance with journal guidelines in advance of publication. Study results will be published in peer-reviewed journals and presented at national and international conferences, community events, to lived experience experts and health advocates, and on social media outlets. In collaboration with a subset of study participants who have completed the study and with NYC H+H/Bellevue providers, we will determine appropriate strategies for communicating study results in both English and Spanish to all study participants, health care providers, and the general public. The study team will identify methodologies and repositories most appropriate for data sharing and management in accordance with the FAIR Guiding Principles (120) and sponsor policies.

## Discussion

Data from this study will confirm or challenge existing paradigms that assert that: 1) long-term pain resolution accompanies surgical weight loss, and 2) reductions in joint mechanical loading are the primary mechanism underlying pain resolution after surgical weight loss. Support for musculoskeletal pain and movement dysfunction after weight loss is limited and often unaddressed in rehabilitation settings(121). Relatedly, there are few recommendations for chronic musculoskeletal pain management beyond the perioperative and acute postsurgical periods in patients undergoing bariatric surgery (42, 122). The results of this study will address critical knowledge gaps in our scientific understanding of the biobehavioral phenotypes of pain following postsurgical weight loss among minoritized populations in the United States. This study has several strengths. First, it employs a comprehensive, standardized battery of tests and measures to assess pain and movement, all of which are scientifically rigorous, reproducible, and clinically relevant. This represents a substantive departure from static, self-reported measures toward a more comprehensive assessment of PAR and MEP, using precise quantification of nociceptive processing and joint motion and loading biosignatures, along with clinical assessment tools commonly used in physical therapist practice. Second, the focus on minoritized U.S. adults highlights salient individual and socioecological factors that often lead to the plateauing or reversal of pain improvements after weight loss(22). Further, this study directly addresses calls to enhance the scientific rigor and trustworthiness of pain research by engaging with populations who bear the greatest pain burden but are often excluded from clinical pain studies(123, 124). Findings from this work could help establish clinically meaningful and community-relevant endpoints and outcomes for future clinical trials. Further, the study results could inform the development and implementation of culturally informed, size-inclusive pain assessments and interventions in a variety of healthcare settings.

There are also limitations. Selection bias refers to systematic factors that can result in the misrepresentation or underrepresentation of the relationship between exposure and outcome(s) in a target study population(125), and can occur at any point from study enrollment to analysis(126). Study attrition among minoritized populations poses a challenge for observational longitudinal studies and may contribute to selection bias. Limited work-schedule flexibility, difficulty finding childcare, long commutes, and strained financial resources significantly hinder retention in weight-loss studies (127). Based on our previous work, we estimate a study attrition rate of 20-40% at the 6-month follow-up (22, 34). We acknowledge potential threats to the study’s internal validity, such as bias and confounding, that may arise directly or indirectly from study attrition (128). The research team will mitigate attrition bias by approaching and screening all eligible patients for bariatric surgery. Since the majority of patients attending the bariatric surgery clinics at both sites are Spanish-speaking or bilingual (fluent in English and Spanish), our research team will communicate with prospective participants in their preferred language for receiving health information. Additionally, we will continually evaluate and implement best practices for recruiting and retaining study participants in weight management studies(129). Measurement bias can arise from systematic errors in data collection and from respondents’ understanding of and willingness to answer survey items, which may influence scoring and the clinical interpretation of the measurement tool (130). To mitigate measurement bias, we will use validated surveys in participants’ preferred language. Additionally, the research team will review participant surveys upon request and will periodically perform data validation to monitor for missing item responses and ensure data quality. A better understanding of the primary driver(s) of CWP dimensions and the key drivers of the intervention response to surgical weight loss could help identify pain responders and non- responders after weight loss who would significantly benefit from adjuvant analgesic or non-pharmacological interventions for continued pain relief during the postsurgical period.

## Data Availability

No datasets were generated or analyzed during the current study. All relevant data from this study will be made available upon study completion.

## Acknowledgements

The authors would like to express their appreciation to the dedicated clinical and administrative staff and the doctors at New York City Health + Hospitals/Bellevue Hospital Center Weight Management and Bariatric Surgery clinics for their assistance with study conceptualization, design, and recruitment. The authors would also like to express their sincerest gratitude to all study participants. ¡Muchas gracias por su tiempo!

