## Supplementary material for "Biobehavioral pain profiling of minoritized adults with chronic widespread pain and clinical obesity before and after bariatric surgery: study protocol for a longitudinal, observational cohort study": Table 1

| **Construct** | **Measure** | **Description** | **B** | **3M** | **6M** | |
| --- | --- | --- | --- | --- | --- | --- |
| **Descriptive Information** | | | | | | |
| Demographic Information* | Standardized electronic forms |  | 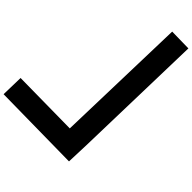 |  | 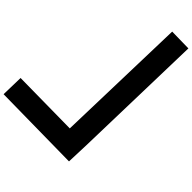 | |
| Age  Sex Assigned at Birth  Gender Identity  Race/ethnicity*  Nationality/Country of Origin  Employment status  Educational Level  Current Address  Household income  English proficiency |  |  |  |  |  | |
| **Anthropometric Measurement** | | | | | | |
| Height (cm) | Stadiometer | Height (cm) will be measured using a stadiometer. | 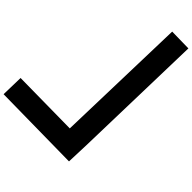 |  | 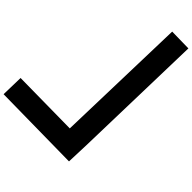 | |
| Weight (kg) | Digitized bariatric scale | Body weight will be measured in clothing using a Stow-A-Weigh digitized bariatric hospital scale. | 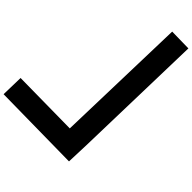 | 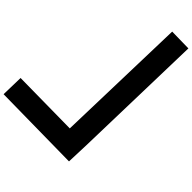 | 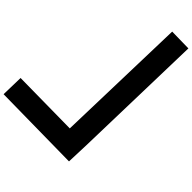 | |
| Body fat distribution | Waist circumference (cm) | A measuring tape is applied at the midpoint between the iliac crest and the lowest rib. | 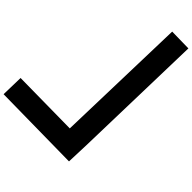 |  | 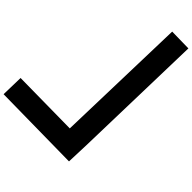 | |
| BMI |  | Weight (kg)/height (m^2^) | 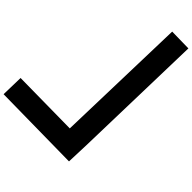 | 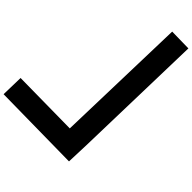 | 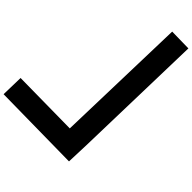 | |
| Weight loss | Percentage of total weight loss (%TWL) | Weight lost after surgery compared to presurgical weight. (Baseline Weight − Current Weight) / ((Baseline Weight) × 100) |  |  | 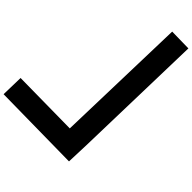 | |
| **Medical History** | | | | | | |
| Bariatric surgery date  Co-morbid health conditions  Current medications | Electronic health records | Surgery type, medical comorbidities, and current medications associated with pain and other health conditions. | 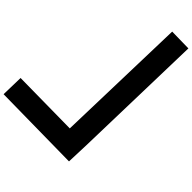 |  | 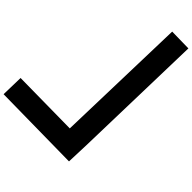 | |
| Sensory Testing | Quantitative Sensory Testing (QST) | Assessment for neuropathy, neuropathic pain, and allodynia. | 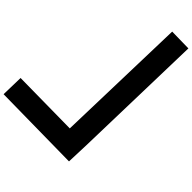 |  | 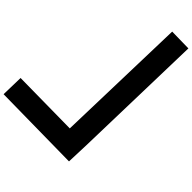 | |
| **Clinical Pain** | | | | | | |
| Pain Intensity at Rest (PAR) | Numeric Pain Rating Scale (NPRS)  BPI- Short Form | The NPRS is a single-item pain intensity scale ranging from 0 (no pain) to 10 (worst pain imaginable). | 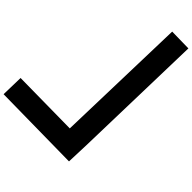 | 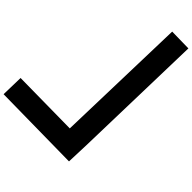 | 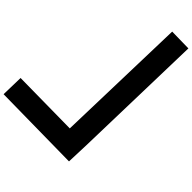 | |
| Movement-Evoked Pain (MEP) | NPRS (0-10) before, during, and after movement tasks | Aggregate pain rating scores across movement tasks divided by the number of tasks. Higher scores indicate greater MEP. | 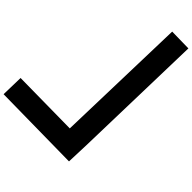 |  | 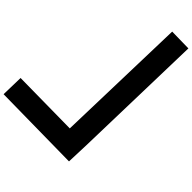 | |
| Pain Location | BPI Short Form (Question 2a) | Assessed via one question (2a), a numerically coded body map. | 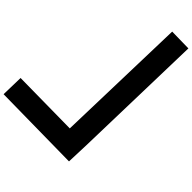 | 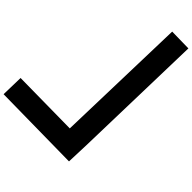 | 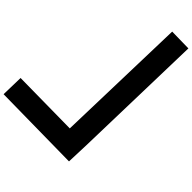 | |
| Pain Chronicity | Self-Report | Participants will answer 1 question about pain duration in months or years | 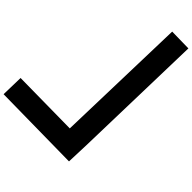 |  |  | |
| Pain Interference | BPI- Short Form | The BPI-SF Pain Severity and Pain Interference subscales feature anchors from 0 (“no pain”) to 10 (“Pain As Bad As You Can Imagine”). The Pain Severity subscale score is the mean of the four items. The BPI-SF Pain Interference subscale is the mean of 7 items. | 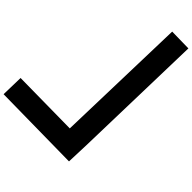 | 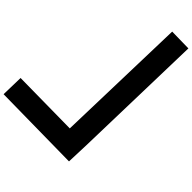 | 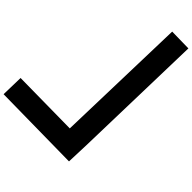 | |
| Pain Behavior | PROMIS SF v.10 – Pain Behavior Short-Form 7a | Items are rated on a 6-point Likert scale, with responses ranging from “Had no pain” to “Always”. Higher T-scores (>50) indicate more pain behaviors. | 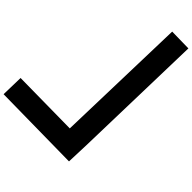 | 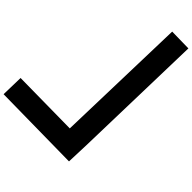 | 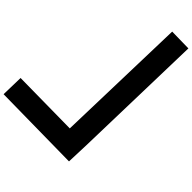 | |
| Central Sensitization | Central Sensitization Inventory (CSI) | The CSI is a 25-item survey that queries how frequently respondents experience symptoms on a Likert scale with anchors from 0 (never) to 4 (always). Total scores range from 0 to 100, with higher scores indicating greater central sensitization. | 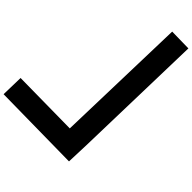 | 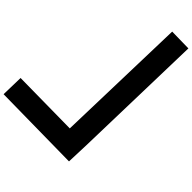 | 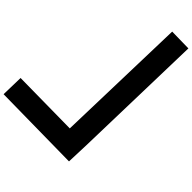 | |
| **Experimental Pain** | | | | | | |
| Quantitative Sensory Testing (QST) |  |  |  |  |  | |
| Static pain sensitivity | Pressure pain threshold (PPT) | Mechanical pressure will be applied using a 1cm^2^ algometer probe at a ramp rate of 50 kPa/sec. PPT will be measured at the upper trapezius, lower back, and medial knee. | 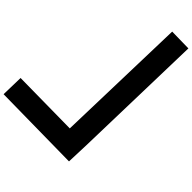 |  | | 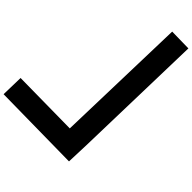 |
|  | Cold Pressor Test (CPT) | Participants will submerge their non-dominant hand up to the wrist crease in a mixture of ice and wateratg8m°- 12 2°C. Participants will rate their pain intensity on a scale from 0 to 10 during and 30 seconds after hand immersion. Pain intensity and immersion time will be recorded. | 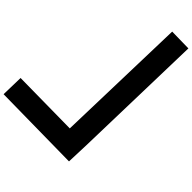 |  | | 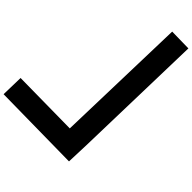 |
| Dynamic pain sensitivity | Mechanical temporal summation (TS) | Participants will rate their pain after a single pin-prick stimulus followed by a 10-train stimulus. A larger absolute effect of TS indicates greater dynamic pain sensitivity. | 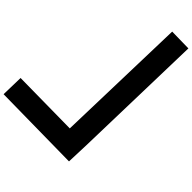 |  | 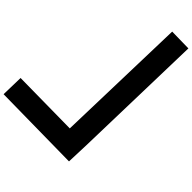 | |
| Descending Pain Inhibition | Conditioned Pain Modulation (CPM) | **Test stimulus:** mechanical pressure **Conditioning stimulus:** cold-water immersion. PPT will be measured immediately before (PPT1) and 60 seconds after (PPT2) cold water immersion. Lower values indicate impaired function of descending inhibitory pathways. | 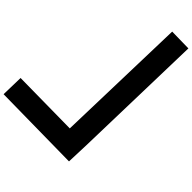 |  | 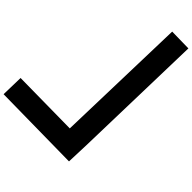 | |
| **Physical Function** | | | | | | |
| Self-reported Function    Performance-Based Function | PROMIS Physical Function Short Form v2 8b | Self-assessment of respondents’ abilities to perform select physical activities Responses range from “Without any difficulty” to “Unable to do on a 5-point Likert scale. Lower T-scores (<50) indicate worse physical function. | 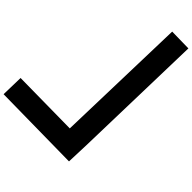 | 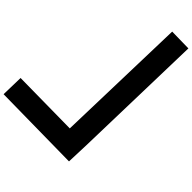 | 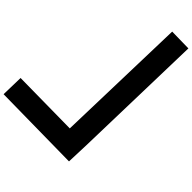 | |
|  | Lower Extremity Functional Scale (LEFS) | Asks about respondents’ difficuly with performing functional activities involving the lower extremities. Total scores range from 0 to 80, with lower scores indicating poorer lower extremity function. | 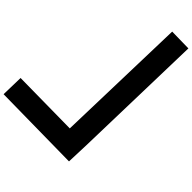 | 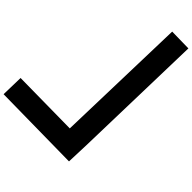 | 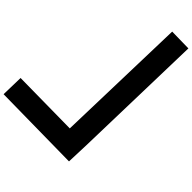 | |
|  | Borg Rate of Perceived Exertion (RPE) | Level of perceived intensity during physical activity. Total scores range from 6-20, with higher scores indicating greater levels of exertion. | 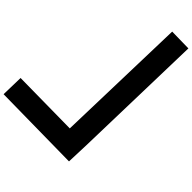 |  | 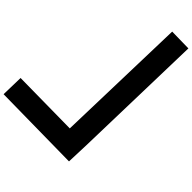 | |
|  | Five Time Sit-to-Stand Test (5TSTS) | Participants will transition from sitting to standing for 5 repetitions. The time (sec) it takes to complete will be recorded and analyzed. | 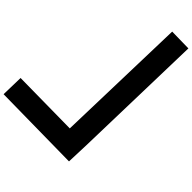 |  | 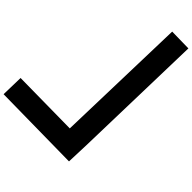 | |
|  | Six-Minute Walk Test (6MWT) | The distance (meters) participants walk for six minutes along a pre-measured 50-foot (15.2 m) walkway. | 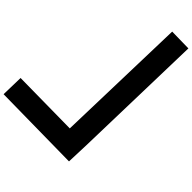 |  | 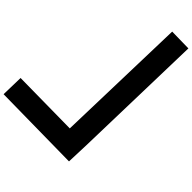 | |
| **Fatigue** | | | | | | |
| Fatigue (at rest, movement-evoked) | NPRS (0-10) before, during, and after movement tasks | Self-reported fatigue intensity using an 11-point rating scale, ranging from 0-10. Higher ratings indicate higher fatigue. | 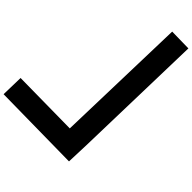 | 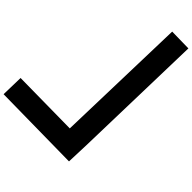 | | 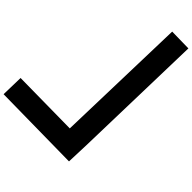 |
|  | PROMIS Short Form v1.0 – Fatigue 13a (FACIT-Fatigue) | Self-reported assessment of the frequency, timing, and severity of fatigue as well as the impact of fatigue on physical, cognitive, and social activities over the past 7 days. Higher T-scores (>50) indicate greater fatigue. | 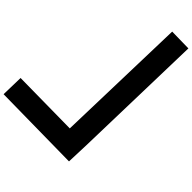 | 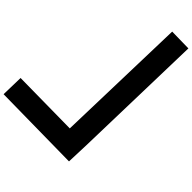 | | 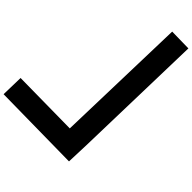 |
| **Psychological, Social, and Emotional Functioning** | | | | | | |
| Pain Catastrophizing | Pain Catastrophizing Scale (PCS) | The Pain Catastrophizing Scale (PCS) is a 13-item self-report instrument. Scores range from 0 to 52, with scores above 30 indicating clinically significant degrees of pain catastrophizing. | 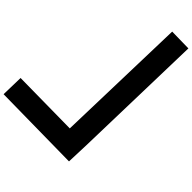 | 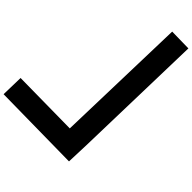 | 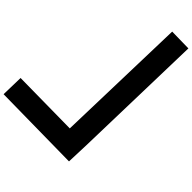 | |
| Perceived Stress | Perceived Stress Scale-10 (PSS-10) | The PSS-10 assesses the extent to which different situations are perceived as stressful. Items are rated on a 5-point Likert scale with anchors from 0 (never) to 4 (very often). Scores are summed and range from 0 to 40. Higher scores indicate greater levels of perceived stress. | 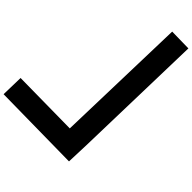 | 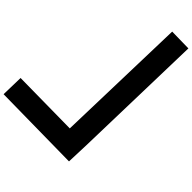 | 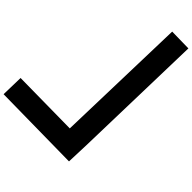 | |
| Depression | PROMIS Short Form v1.0 - Depression 8a | Items are rated on a 5-point Likert scale, with responses ranging from “Never” to “Always”. Items are summed to yield a total raw score, which is then converted to a standardized T-score. Higher T-scores (<50) indicate higher levels of depression. | 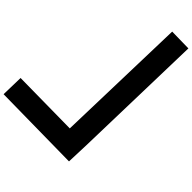 | 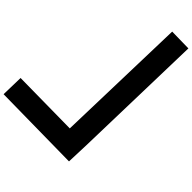 | 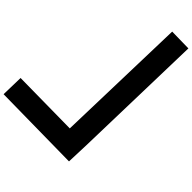 | |
| Emotional Support | PROMIS SF v2.0 - Emotional Support 6a questionnaire | Items are rated on a 5-point Likert scale, with responses ranging from “Never” to “Always”. Items are summed to yield a total raw score, which is then converted to a standardized T-score. Lower T-scores (<50) indicate lower levels of emotional support. | 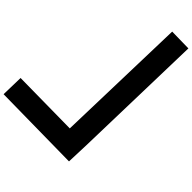 | 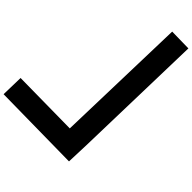 | 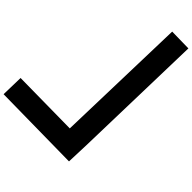 | |
| Sleep Quality | Pittsburgh Sleep Quality Index (PSQI) | The PSQI survey assesses sleep functioning over the last month. Items are rated on a 4-point Likert scale with anchors from 0 (no difficulty) to 3 (severe difficulty). The total score ranges from 0 to 21. Higher scores greater than 5 indicate poor sleep quality. | 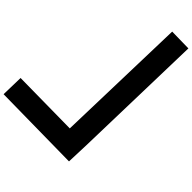 | 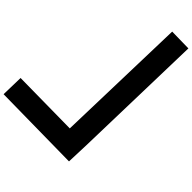 | 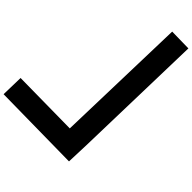 | |
| **Joint Motion and Muscle Strength** | | | | | | |
| Clinical measures of joint motion  (knee, thoracolumbar) | Goniometry  Gravity-dependent bubble inclinometry | **Goniometry:** Active knee flexion and extension will be measured in the supine position, using a universal goniometer. **Inclinometry:** Active flexion and extension of the thoracic and lumbar spine will be measured. | 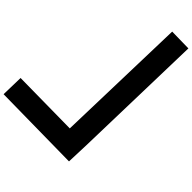 |  | | 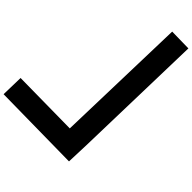 |
| Instrumented measures of joint  motion and mechanical loading  (trunk, hip, knee, ankle) | Marker-based motion capture (stereophotogrammetry) | Quantifies the three-dimensional positions and orientations of the trunk, pelvis, thigh, shank (leg), and foot segments during select movement tasks. | 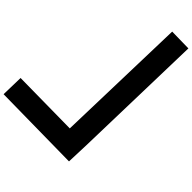 |  | | 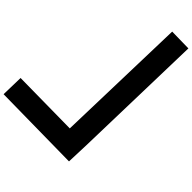 |
| Muscle strength | Handheld dynamometry (HHD) | A tester will administer the “break test,” in which a perpendicular force is applied at the end of knee extension bilaterally. | 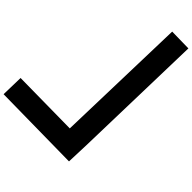 |  | 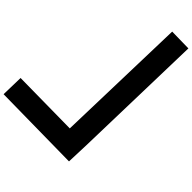 | |
